# Factors associated with newly diagnosed ischemic stroke among people in Thailand: A population-based case-control study

**DOI:** 10.1101/2025.01.29.25321321

**Authors:** Junjira Phasom, Ratchadaporn Ungcharoen, Pakorn Pusuwan, Nitikorn Phoosuwan

## Abstract

**Background:** Stroke is a leading cause of death and disability-adjusted life years globally. The incidence of stroke is increasing in Asia, with ischemic stroke accounting for approximately 80% of stroke cases in Thailand. Stroke often results in long-term disabilities, including impairments in speech, communication, and concentration.

**Objective:** This study aimed to investigate factors associated with newly diagnosed ischemic stroke.

**Methods:** A matched case-control study was conducted, including 154 newly diagnosed ischemic stroke cases and 183 non-stroke individuals with type 2 diabetes mellitus (T2DM) as controls. Data were collected between February and September 2022 (post-COVID-19 period) using a structured questionnaire covering (1) socio-demographics, (2) lifestyle factors, (3) perceived social support, and (4) self-care management (SCM). Multivariable logistic regression models were employed to calculate adjusted odds ratios (aOR) with 95% confidence intervals (CI).

**Results:** Most participants were female (60.8%), Buddhists (92.9%), and agriculturists (66.5%), with a mean age of 58.9 (±9.9) years. Factors associated with ischemic stroke included male sex (aOR=3.533, 95%CI 1.732–7.206), Buddhism (aOR=3.529, 95%CI 1.107–11.250), sedentary occupation (aOR=5.785, 95%CI 2.613–12.807), and having T2DM for over 10 years (aOR=6.194, 95%CI 3.553–10.798). Protective factors included age ≥ 60 years (aOR=0.553, 95%CI 0.312–0.982) and moderate SCM levels (aOR=0.453, 95%CI 0.256–0.802).

**Conclusion:** Prolonged T2DM and sedentary occupations significantly contribute to ischemic stroke incidence. Effective prevention programs, including lifestyle modifications and diabetes self-care management education, may help reduce the burden of ischemic stroke.

## Introduction

Stroke is the second leading cause of death globally and the third leading cause of combined death and disability, with its burden increasing rapidly in low-income (LICs) and middle-income countries (MICs) [1]. Between 1990 and 2019, the global burden of stroke increased significantly, with incidence rising by 70% (12.2 million new cases), prevalence by 85% (101 million cases), mortality by 43% (6.55 million deaths), and disability-adjusted life years (DALYs) by 32% (143 million DALYs) [2]. In Asia, the incidence rate of stroke ranges from 116 to 483 per 100,000 population per year [3]. Ischemic stroke, defined necrosis of brain tissue caused by the narrowing or blockage of arteries supplying blood to the brain, leads to insufficient blood flow and consequent brain damage [4]. It accounts for approximately 62% of all stroke cases, with the remaining 28% classified as hemorrhagic strokes [5]. Stroke often results in chronic conditions or long-term disabilities, such as post-stroke cognitive impairment (PSCI) and post-stroke dementia (PSD) [6, 7], as well as significant physical impairments [8–10].

Globally, ischemic stroke accounts for 7.6 million new cases annually, with over 58% occurring in individuals under the age of 70. It is responsible for 3.3 million deaths and over 63 million DALYs each year [5]. Between 1990 and 2019, the age-standardized incidence rate of ischemic stroke increased from 96.09 to 104.89 per 100,000 population, while the age-standardized mortality rate declined from 60.98 to 48.69 per 100,000 population [4].

Risk factors for stroke are classified as non-modifiable, semi-modifiable, and modifiable.

Non-modifiable risk factors include gender, age [6, 11], and family history [12]. Semi- modifiable factors include hypertension, diabetes, hypercholesterolemia/dyslipidemia [13, 14], and air quality [2, 15]. Modifiable factors include smoking, alcohol consumption, physical inactivity, and obesity [2, 15].

Thailand, classified as an upper-MIC [16], is transitioning into an aged society [17]. The burden of noncommunicable diseases has increased, as evidenced by rising cases of type 2 diabetes mellitus (T2DM), hypertension, and stroke [18]. Stroke remains a major health challenge in Thailand and is the leading cause of death, with a higher prevalence observed in males than females [19]. The overall prevalence of stroke in Thailand is 1.3% [6], with ischemic stroke accounting for approximately 80% of all stroke cases and hemorrhagic stroke comprising the remaining 20% [19].

A review of the literature revealed only eight studies investigating risk factors for stroke in Thailand. These include four nationwide studies, three focused on the central region, and one conducted in the northeast region. However, these studies did not examine ischemic stroke specifically or consider regional variations [19–22]. Therefore, this study was aimed to investigate the factors associated with newly diagnosed ischemic stroke in Thailand. The findings may inform the development of targeted interventions to prevent ischemic stroke.

## Materials and Methods

### Aim

This matched case-control study was aimed to investigate factors associated with newly diagnosed ischemic stroke in Thailand.

### Setting

This study was conducted in Sakon Nakhon, a northeastern province of Thailand with a population of approximately 1.2 million and an annual per capita income of around 2,100 USD [23]. The incidence rate of stroke in Sakon Nakhon was approximately 295.4 per 100,000 population per year [24].

### Participants

The sample size was calculated using a multiple logistic regression formula [25] with an odds ratio of 2.8 [14], a confidence level of 95% (α = 0.05), a power of 80% (ß = 0.8), and a case-to-control ratio of 1:1. The minimum sample size required for both cases and controls was 150 participants each. Inclusion criteria for cases included individuals who: (1) were admitted to the stroke unit at Sakon Nakhon Provincial Hospital with a clinical diagnosis of first-time ischemic stroke (ICD-10 codes I630–I639 within seven days of onset), (2) were of Thai ethnicity, (3) could communicate in Thai, and (4) were aged between 18 and 75 years. Inclusion criteria for controls included individuals who: (1) were recruited from communities similar to those where the cases resided (living in the community for over one year), (2) had never been diagnosed with ischemic stroke (ICD-10 codes I630–I639), (3) matched the cases in gender and age group, (4) could communicate in Thai, and (5) had a history of diabetes for more than one year. Exclusion criteria for both cases and controls included individuals who: (1) declined to participate in the study or (2) exhibited signs of cognitive impairment.

### Procedure

The study protocol was submitted to and approved by ethics committees in Thailand. Researchers contacted the head nurse of the stroke unit to identify and recruit cases and collaborated with the directors of Subdistrict Health Promoting Hospitals (SHPHs) in the same areas as the cases to recruit controls. A total of 154 out of 185 newly diagnosed ischemic stroke cases (response rate: 83.2%) and 183 out of 185 non-stroke individuals with type 2 diabetes mellitus (T2DM) as controls (response rate: 98.9%) participated. Data collection was conducted between February 21^st^, and September 30^th^, 2022, which was during the post-COVID-19 period.

### Ethical approval

Ethical approval was obtained from the Sakon Nakhon Provincial Hospital Ethics Committee (COA026/2564) and Kasetsart University Chalermphrakiat Sakon Nakhon Province Campus Ethics Committee (Kucsc.HE-64-008). The study adhered to the ethical principles outlined in the Declaration of Helsinki. Permissions were also obtained from the director of Sakon Nakhon Provincial Hospital, the head nurse of the stroke unit, and the directors of the selected SHPHs.

Prior to participation, all participants were provided with both oral and written information about the study. They signed informed consent forms voluntarily at the stroke unit (for cases) and at a designated room near the selected SHPHs (for controls). Participants were informed of their right to withdraw from the study at any time without consequences. Confidentiality was strictly maintained, and participant data were anonymized throughout the study.

### Measurement

A structured questionnaire was used for both cases and controls, comprising five sections: socio-demographics, the Alcohol Use Disorders Identification Test (AUDIT), self-care management (SCM), the Revised Thai Multi-Dimensional Scale of Perceived Social Support (r-T-MSPSS), and the Fagerström Test for Nicotine Dependence (FTND). The questionnaire was reviewed for content validity by three experts and pretested for reliability among individuals in a community near Sakon Nakhon. Both content validity and reliability scores were deemed acceptable.

The socio-demographic section included information on gender, age, marital status, education level, occupation, family history of paralysis, family history of cardiovascular disease, smoking status, alcohol consumption, and duration of T2DM.

The AUDIT, developed by the World Health Organization [26], was used to assess alcohol consumption, potential dependence, and alcohol-related harm. It consisted of 10 items scored from 0 to 4, except for questions nine and ten, which were scored as 0, 2, or 4. Total scores were categorized as follows: 1–7 (low-risk consumption), 8–14 (hazardous or harmful consumption), and 15 or more (indicating moderate to severe alcohol use disorder). The Thai version of the AUDIT, translated by the Thai Health Promotion Foundation [27], was used in this study.

The SCM section, developed by Han et al. [28], measured self-care behaviors, motivation, and self-efficacy, based on Orem’s self-care model and Motivational Interviewing. The instrument comprised 20 items, each rated on a 6-point scale (1–6), with total scores ranging from 20 to 120. The total scores were categorized using Bloom’s classification [29]: 20–47 (low SCM), 48–63 (moderate SCM), and 64–80 (high SCM). The researchers translated the English version into Thai using a backward-forward translation process.

The r-T-MSPSS, originally developed by Zimet et al. [30] and translated into Thai by Wongpakaran et al. [31], assessed perceived social support. It consisted of 12 items scored on a 7-point Likert scale (1 = very strongly disagree, 7 = very strongly agree), with total scores ranging from 12 to 84. Scores were categorized as follows: 12–36 (low perceived social support), 37–60 (moderate perceived social support), and 61–84 (high perceived social support) [32].

The FTND, modified by Heatherton et al. [33] and translated into Thai by Klinsophon et al. [34], assessed nicotine dependence using six items, each scored from 0 to 3. Total scores were categorized as follows: 1–2 (very low dependence), 3–4 (low dependence), 5 (moderate dependence), 6–7 (high dependence), and 8 or more (very high dependence).

### Data Analysis

Descriptive statistics, including frequency, mean, standard deviation (SD), minimum, maximum, and percentage (%), were used to summarize the data. Inferential statistics were used to explore associations between independent and dependent variables using multivariable logistic regression. Cross-tabulations and Chi-square tests were used to analyze relationships between categorical variables, with p-values indicating significance. Univariable logistic regression was conducted to assess individual relationships between variables and ischemic stroke. Variables with p ≤ 0.05 in univariable regression were included in multivariable regression analysis. Crude odds ratios (cOR), adjusted odds ratios (aOR), and 95% confidence intervals (CI) were calculated to examine the associations between risk factors and ischemic stroke. Statistical significance was set at p < 0.05, and all tests were two-sided. The independent variables included socio-demographic factors (sex, age, marital status, education level, occupation, family history of paralysis, family history of cardiovascular disease, smoking, and alcohol consumption), SCM, and r-T-MSPSS. The dependent variable was ischemic stroke.

## Results

A total of 337 participants were included in the study, comprising 154 cases and 183 controls. The majority of participants were female (60.8%), and the mean age was 58.9 years (±9.9). Most participants were Buddhists (92.9%), married (78.0%), had a primary education or less (76.3%), worked as agriculturists (66.5%), never smoked (73.6%), never consumed alcohol (61.4%), had no family history of paralysis (92.6%), and had no family history of cardiovascular disease (97.3%) (Table 1).

**Table 1.** Characteristics of participants.

| Characteristics | Total | Cases | Controls | $\chi^2$ | p-value |
| --- | --- | --- | --- | --- | --- |
|  | n (%) | n (%) | n (%) |  |  |
| **Overall** | 337 | 154 (45.7) | 183 (54.3) |  |  |
| **Sex** |  |  |  | 38.451 | <0.001* |
| Male | 132 (39.2) | 88 (57.1) | 44 (24.0) |  |  |
| Female | 205 (60.8) | 66 (42.9) | 139 (76.0) |  |  |
| **Religion** |  |  |  | 6.438 | 0.011* |
| Buddhism | 313 (92.9) | 149 (96.8) | 164 (89.6) |  |  |
| Christianity | 24 (7.1) | 5 (3.2) | 19 (10.4) |  |  |
| **Age (years)** |  |  |  | 1.049 | 0.592 |
| ≤50 | 59 (17.5) | 30 (19.5) | 29 (15.9) |  |  |
| 51-60 | 117 (34.7) | 50 (32.5) | 67 (36.6) |  |  |
| ≥61 | 161 (47.8) | 74 (48.0) | 87 (47.5) |  |  |
| Mean ± SD | 58.9 ± 9.9 | 58.5 ± 11.1 | 59.3 ± 8.9 |  |  |
| (Min, Max) | (21, 75) | (21, 75) | (28, 75) |  |  |
| **Marital status** |  |  |  | 11.812 | 0.008* |
| Single | 15 (4.5) | 4 (2.6) | 11 (6.0) |  |  |
| Married | 263 (78.0) | 123 (79.9) | 140 (76.5) |  |  |
| Divorced | 44 (13.0) | 15 (9.7) | 29 (15.9) |  |  |
| Widowed | 15 (4.5) | 12 (7.8) | 3 (1.6) |  |  |
| **Education level** |  |  |  | 5.745 | 0.057 |
| Primary school or less | 257 (76.3) | 112 (72.7) | 145 (79.2) |  |  |
|  | n (%) | n (%) | n (%) |  |  |
| Secondary school | 57 (16.9) | 26 (16.9) | 31 (17.0) |  |  |
| Higher education | 23 (6.8) | 16 (10.4) | 7 (3.8) |  |  |
| **Occupation** |  |  |  | 45.494 | <0.001* |
| Agriculturists | 224 (66.5) | 76 (49.4) | 148 (80.8) |  |  |
| Daily workers | 33 (9.8) | 22 (14.3) | 11 (6.0) |  |  |
| Self employers | 20 (5.9) | 12 (7.8) | 8 (4.4) |  |  |
| Civil servants | 19 (5.6) | 13 (8.4) | 6 (3.3) |  |  |
| Unemployed | 31 (9.2) | 27 (17.5) | 4 (2.2) |  |  |
| Housewives | 10 (3.0) | 4 (2.6) | 6 (3.3) |  |  |
| **Family history of paralysis** |  |  |  | 0.031 | 0.859 |
| No | 312 (92.6) | 143 (92.9) | 169 (92.3) |  |  |
| Yes | 25 (7.4) | 11 (7.1) | 14 (7.7) |  |  |
| **Family history of cardiovascular disease** |  |  |  | 0.361 | 0.547 |
| No | 328 (97.3) | 149 (96.8) | 179 (97.8) |  |  |
| Yes | 9 (2.7) | 5 (3.2) | 4 (2.2) |  |  |
| **Smoking** |  |  |  | 26.179 | <0.001* |
| Never smoked | 248 (73.6) | 93 (60.4) | 155 (84.7) |  |  |
| Quit smoking | 35 (10.4) | 22 (14.3) | 13 (7.1) |  |  |
| Current smoker | 54 (16.0) | 39 (25.3) | 15 (8.2) |  |  |
|  | n (%) | n (%) | n (%) |  |  |
| **FTND level** |  |  |  | 20.322 | <0.001* |
| Never smoked or quit smoking | 283 (84.0) | 115 (74.7) | 168 (91.8) |  |  |
| High dependence (6-7 points) | 20 (5.9) | 17 (11.0) | 3 (1.6) |  |  |
| Very high dependence ($\geq 8$ points) | 34 (10.1) | 22 (14.3) | 12 (6.6) | | |
| **Alcohol Consumption** |  |  |  | 17.453 | <0.001* |
| Never had alcohol | 207 (61.4) | 76 (49.4) | 131 (71.6) |  |  |
| Quit alcohol | 47 (14.0) | 28 (18.2) | 19 (10.4) |  |  |
| Currently take alcohol | 83 (24.6) | 50 (32.4) | 33 (18.0) |  |  |
| **AUDIT level** |  |  |  | 3.574 | 0.167 |
| Never had alcohol or Quit alcohol (0 point) | 254 (75.4) | 123 (79.9) | 131 (71.6) |  |  |
| Low-risk consumption (1-7 points) | 70 (20.8) | 25 (16.2) | 45 (24.6) |  |  |
| Harmful alcohol consumption ($\geq 8$ points) | 13 (3.8) | 6 (3.9) | 7 (3.8) | | |
| **SCM Level** |  |  |  | 8.890 | 0.012* |
| Low (20-47 points) | 123 (36.5) | 69 (44.8) | 54 (29.5) |  |  |
| Moderate (48-63 points) | 201 (59.6) | 81 (52.6) | 120 (65.6) |  |  |
|  | n (%) | n (%) | n (%) |  |  |
| Hight (64-80 points) | 13 (3.9) | 4 (2.6) | 9 (4.9) |  |  |
| Mean ± SD | 50.3 ± 7.9 | 48.9 ± 7.8 | 51.6 ± 7.7 | | |
| (Min, Max) | (30, 72) | (30, 69) | (33, 72) |  |  |
| **r-T-MSPSS Level** |  |  |  | 3.082 | 0.214 |
| Low (12-36 points) | 12 (3.6) | 3 (2.0) | 9 (4.9) |  |  |
| Moderate (37-60 points) | 110 (32.6) | 47 (30.5) | 63 (34.4) |  |  |
| High (61-84 points) | 215 (63.8) | 104 (67.5) | 111 (60.7) |  |  |
| Mean ± SD | 64.6 ± 13.2 | 66.0 ± 12.7 | 63.5 ± 13.5 | | |
| (Min, Max) | (16, 84) | (36, 84) | (16, 84) |  |  |
| **Duration of T2DM** |  |  |  | 59.730 | <0.001* |
| $\leq 10$ years | 180 (53.4) | 47 (30.5) | 133 (72.7) | | |
| $\geq 11$ years | 157 (46.6) | 107 (69.5) | 50 (27.3) | | |
| Mean ± SD | 8.6 ± 7.5 | 9.9 ± 9.4 | 8.1 ± 6.7 | | |
| (Min, Max) | (1, 40) | (1, 40) | (1, 30) |  |  |

Abbreviations: SD, standard deviation; AUDIT, Alcohol Use Disorders Identification Test; FTND, Fagerström Test for Nicotine Dependence; SCM, Self-care management; r-T-MSPSS, The Revised Multi-Dimensional Scale of Perceived Social Support; T2DM, type 2 diabetes mellitus; \*Significance level of $p < 0.05$

Factors significantly associated with newly diagnosed ischemic stroke included being male (aOR = 3.533, 95% CI: 1.732–7.206), practicing Buddhism (aOR = 3.529, 95% CI: 1.107– 11.250), having a sedentary occupation (aOR = 5.785, 95% CI: 2.613–12.807), and having type 2 diabetes mellitus (T2DM) for more than 10 years (aOR = 6.194, 95% CI: 3.553–10.798).

Protective factors included being aged 60 years or older (aOR = 0.553, 95% CI: 0.312–0.982) and having a moderate level of self-care management (SCM) (aOR = 0.453, 95% CI: 0.256– 0.802) (Table 2, Fig. 1).

**Figure 1.**
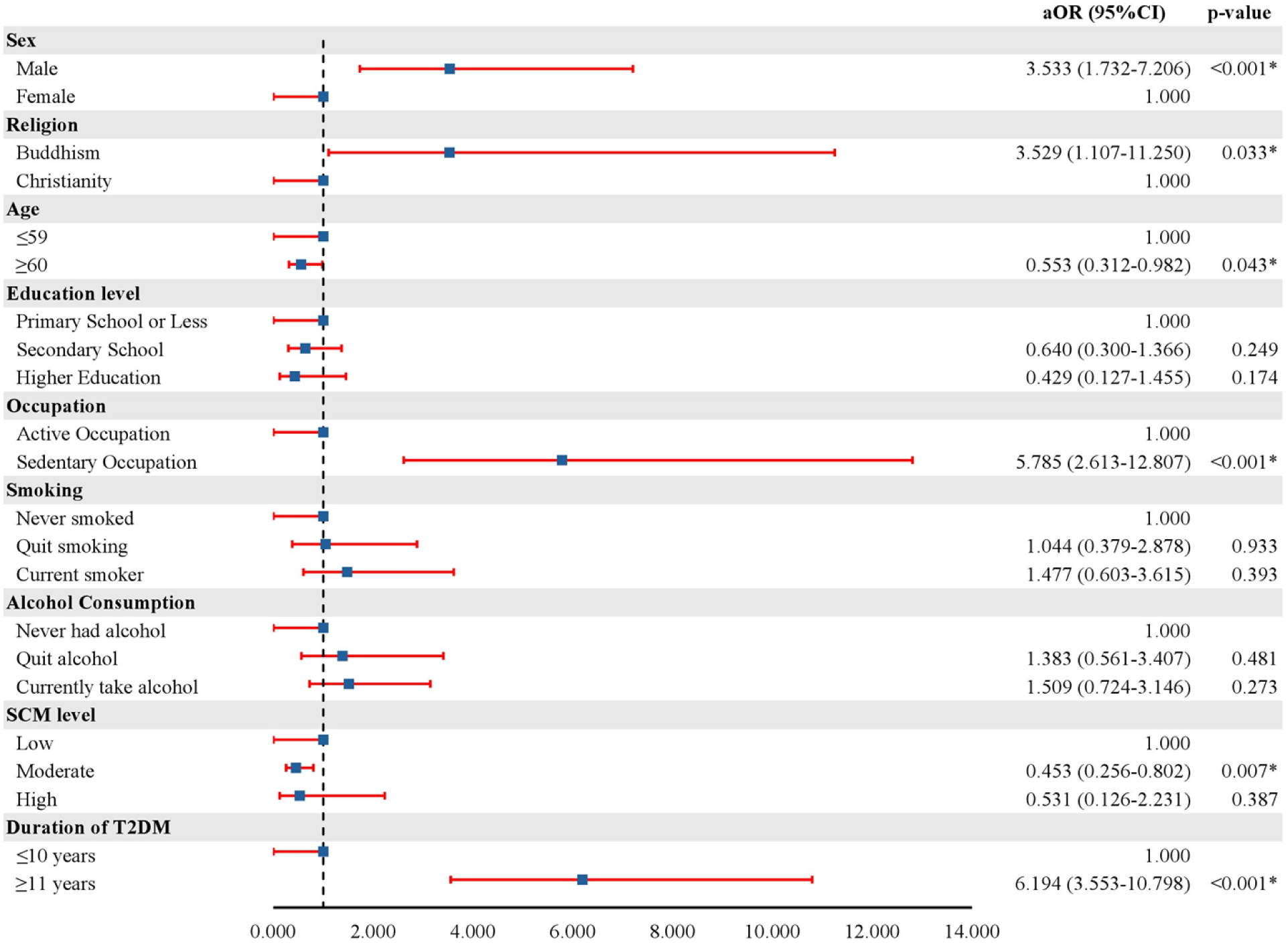
Forest plot displaying the adjusted odds ratios (aOR) and 95% confidence intervals (CI) for ischemic stroke risk factors. Abbreviations: aOR, adjusted odds ratios; CI, confidence intervals; SCM, self-care management; T2DM, type 2 diabetes mellitus; *Significance level of p < 0.05.

**Table 2.** Univariable and multivariable analyses of participant’s characteristics of ischemic stroke.

| Characteristics | Ischemic Stroke |  | cOR (95%CI) | p-value | aOR (95%CI) | p-value |
| --- | --- | --- | --- | --- | --- | --- |
|  | Yes | No |  |  |  |  |
|  | n (%) | n (%) |  |  |  |  |
| **Overall** | 154 (45.7) | 183 (54.3) |  |  |  |  |
| **Sex** |  |  |  |  |  |  |
| Male | 88 (57.1) | 44 (24.0) | 4.212 (2.644-6.710) | <0.001* | 3.533 (1.732-7.206) | <0.001* |
| Female | 66 (42.9) | 139 (76.0) | 1.000 |  | 1.000 |  |
| **Religion** |  |  |  |  |  |  |
| Buddhism | 149 (96.8) | 164 (89.6) | 3.452 (1.258-9.477) | 0.016* | 3.529 (1.107-11.250) | 0.033* |
| Christianity | 5 (3.2) | 19 (10.4) | 1.000 |  | 1.000 |  |
| **Age (years)** |  |  |  |  |  |  |
| ≤59 | 78 (50.6) | 92 (50.3) | 1.000 |  | 1.000 |  |
| ≥60 | 76 (49.4) | 91 (49.7) | 0.985 (0.642-1.512) | 0.945 | 0.553 (0.312-0.982) | 0.043* |
| **Marital status** |  |  |  |  |  |  |
| Never Married | 4 (2.6) | 11 (6.0) | 1.000 |  |  |  |
| Ever Married | 150 (97.4) | 172 (94.0) | 2.398 (0.748-7.690) | 0.141 |  |  |
|  | Yes | No |  |  |  |  |
|  | n (%) | n (%) |  |  |  |  |
| **Education level** |  |  |  |  |  |  |
| Primary school or less | 112 (72.7) | 145 (79.2) | 1.000 |  | 1.000 |  |
| Secondary school | 26 (16.9) | 31 (17.0) | 1.086 (0.610-1.933) | 0.780 | 0.640 (0.300-1.366) | 0.249 |
| Higher Education | 16 (10.4) | 7 (3.8) | 2.959 (1.177-7.439) | 0.021* | 0.429 (0.127-1.455) | 0.174 |
| **Occupation** |  |  |  |  |  |  |
| Active occupation | 102 (66.2) | 165 (90.2) | 1.000 |  | 1.000 |  |
| Sedentary occupation | 52 (33.8) | 18 (9.8) | 4.673 (2.590-8.431) | <0.001* | 5.785 (2.613-12.807) | <0.001* |
| **Family history of paralysis** |  |  |  |  |  |  |
| No | 143 (92.9) | 169 (92.3) | 1.000 |  |  |  |
| Yes | 11 (7.1) | 14 (7.7) | 0.929 (0.409-2.109) | 0.859 |  |  |
| **Family history of cardiovascular disease** |  |  |  |  |  |  |
|  | Yes | No |  |  |  |  |
|  | n (%) | n (%) |  |  |  |  |
| No | 149 (96.8) | 179 (97.8) | 1.000 |  |  |  |
| Yes | 5 (3.2) | 4 (2.2) | 1.502 (0.396-5.693) | 0.550 |  |  |
| **Smoking** |  |  |  |  |  |  |
| Never smoked | 93 (60.4) | 155 (84.7) | 1.000 |  | 1.000 |  |
| Quit smoking | 22 (14.3) | 13 (7.1) | 2.821 (1.356-5.866) | 0.006* | 1.044 (0.379-2.878) | 0.933 |
| Current smoker | 39 (25.3) | 15 (8.2) | 4.333 (2.265-8.289) | <0.001* | 1.477 (0.603-3.615) | 0.393 |
| **Alcohol Consumption** |  |  |  |  |  |  |
| Never had alcohol | 76 (49.4) | 131 (71.6) | 1.000 |  | 1.000 |  |
| Quit alcohol | 28 (18.2) | 19 (10.4) | 2.540 (1.329-4.854) | 0.005* | 1.383 (0.561-3.407) | 0.481 |
| Currently take alcohol | 50 (32.5) | 33 (18.0) | 2.612 (1.549-4.404) | <0.001* | 1.509 (0.724-3.146) | 0.273 |
| **SCM Level** |  |  |  |  |  |  |
| Low (20-47 points) | 69 (44.8) | 54 (29.5) | 1.000 |  | 1.000 |  |
| Moderate (48-63 points) | 81 (52.6) | 120 (65.6) | 0.528 (0.335-0.832) | 0.006* | 0.453 (0.256-0.802) | 0.007* |
|  | Yes | No |  |  |  |  |
|  | n (%) | n (%) |  |  |  |  |
| Hight (64-80 points) | 4 (2.6) | 9 (4.9) | 0.348 (0.102-1.191) | 0.093 | 0.531 (0.126-2.231) | 0.387 |
| **r-T-MSPSS Level** |  |  |  |  |  |  |
| Low (12-36 points) | 3 (2.0) | 9 (4.9) | 1.000 |  |  |  |
| Moderate (37-60 points) | 47 (30.5) | 63 (34.4) | 2.238 (0.574-8.721) | 0.246 |  |  |
| High (61-84 points) | 104 (67.5) | 111 (60.7) | 2.811 (0.741-10.668) | 0.129 |  |  |
| **Duration of T2DM** |  |  |  |  |  |  |
| ≤10 years | 47 (30.5) | 133 (72.7) | 1.000 |  | 1.000 |  |
| ≥11 years | 107 (69.5) | 50 (27.3) | 6.056 (3.775-9.714) | <0.001* | 6.194 (3.553-10.798) | <0.001* |

Abbreviations: cOR, crude odds ratios; aOR, adjusted odds ratios; CI, confidence intervals; SCM, Self-care management; r-T-MSPSS, The Revised Multi-Dimensional Scale of Perceived Social Support; T2DM, type 2 diabetes mellitus; \*Significance level of $p < 0.05$

## Discussion

This study aimed to investigate factors associated with people newly diagnosed with ischemic stroke in Thailand. The results identified non-modifiable factors (gender, duration of T2DM, and age), semi-modifiable factors (Buddhism and sedentary occupation), and modifiable factors (self-care management [SCM]) as significantly associated with ischemic stroke. The mean age of ischemic stroke cases in this study was 58.5 (±11.1) years, with the majority of participants aged 60 years or younger (52.0%), which aligns with findings from previous studies [20].

Approximately 60% of participants were male, consistent with other studies [20–22]. Furthermore, most participants were married and had a primary level of education, which is also consistent with prior research [20, 22]. These demographic similarities suggest that the findings of this study could be generalized to other contexts.

Prolonged T2DM (11 years or more) was identified as a significant risk factor for ischemic stroke. Hyperglycemia and vascular inflammation associated with T2DM contribute to atherosclerosis and other vascular complications, thereby elevating stroke risk [11]. Previous studies have also demonstrated that diabetes duration significantly increases stroke risk. For instance, individuals with diabetes for over 10 years face a threefold higher risk of ischemic stroke, with an annual increase of 3% [35]. Similarly, a diabetes duration of eight years or more is associated with an elevated risk of 1-year stroke recurrence following ischemic stroke or transient ischemic attack [36]. Additional studies have linked prolonged diabetes with higher stroke risk in patients with atrial fibrillation, especially when accompanied by poor glycemic control (HbA1c >7%) [37, 38]. These findings underscore the importance of screening for ischemic stroke in individuals with prolonged T2DM.

This study also found that having a sedentary occupation significantly increased the risk of ischemic stroke. Sedentary occupations involve minimal physical activity and include professions such as civil servants, self-employed individuals, and the unemployed. Prolonged sedentary behavior, such as sitting for more than eight hours daily, has been associated with an increased risk of ischemic stroke [39]. Although individuals in sedentary occupations may not face high physical workloads, they are often exposed to other risk factors, including reduced physical activity, environmental stressors, and psychosocial pressures, which may contribute to stroke risk [20]. Further research is needed to determine whether the relationship between sedentary occupation and ischemic stroke is causal or influenced by confounding variables.

Being male was also identified as a significant risk factor for ischemic stroke. Both biological and sociocultural factors contribute to this disparity. Sex hormones, such as testosterone and estrogen, play a role in stroke pathophysiology, while sociocultural factors, such as higher rates of smoking and alcohol consumption among males, further elevate their risk [40–42]. Efforts to reduce stroke risk in males should focus on discouraging smoking and excessive alcohol consumption.

## Strengths and Limitations

This study utilized a case-control design with a 1:1 matching ratio for gender, age, and residential area, which is suitable for investigating a disease with a long duration, such as T2DM and stroke. The use of multivariable logistic regression helped adjust for potential confounding factors, enhancing the validity of the findings. Additionally, data collection during the post-COVID-19 period allowed for the exploration of risk factors during a unique global crisis.

However, several limitations should be noted. The reliance on self-reported data may have introduced recall bias, and stroke cases managed outside hospital settings were excluded, potentially introducing selection bias. Furthermore, the study did not account for stroke severity, which may influence the observed associations. Future research should aim to address these limitations by including more diverse samples and exploring causal relationships between identified risk factors and ischemic stroke. A qualitative study focusing on individuals with ischemic stroke may also provide deeper insights into additional risk factors.

## Conclusion

Prolonged diabetes and sedentary occupations are significant contributors to ischemic stroke incidence. Moderate levels of self-care management, including adherence to medical appointments and medications, as well as lifestyle adjustments, may act as protective factors. Additionally, older age (≥60 years) appears to confer some protective effect. Community-based prevention programs, such as diabetes self-care management education, could empower individuals to adopt healthier lifestyles. Early intervention targeting at-risk populations, particularly those with prolonged diabetes or sedentary lifestyles, is essential to reduce the incidence of ischemic stroke.

## Data Availability

All relevant data are within the manuscript and its Supporting Information files.

## Acknowledgements

This research was funded by Kasetsart University through the Graduate School Fellowship Program. The researchers would like to express their gratitude to all participants who contributed their time and effort to this study. Special thanks are extended to the healthcare staff who provided valuable assistance during the data collection process.

## Notes

### Competing Interest Statement

The authors have declared no competing interest.

### Funding Statement

The author(s) received no specific funding for this work.

### Author Declarations

Ethical approval was obtained from the Sakon Nakhon Provincial Hospital Ethics Committee (COA026/2564) and Kasetsart University Chalermphrakiat Sakon Nakhon Province Campus Ethics Committee (Kucsc.HE-64-008).

